# Single-cell RNA sequencing reveals the distinctive roles of CD4^+^ and CD8^+^ T cells in autoimmune uveitis

**DOI:** 10.1101/2022.11.28.22282728

**Authors:** Hao Kang, Hongjian Sun, Yang Yang, Minglei Shu, Yunbo Wei, Yu Zhang, Di Yu, Yong Tao

## Abstract

Uveitis is a common cause of blindness and classified as infectious or non-infectious types. Non-infectious uveitis is often secondary to systemic autoimmune diseases, with Behçet’s disease (BD) and Vogt-Koyanagi-Harada disease (VKHD) as the two most common causes. Uveitis in BD and VKHD often show similar clinical manifestations, but the underlying immunopathogenesis remains unclear. We collected immune cell infiltrates in aqueous humour of BD and VKHD uveitis patients and analysed them by single-cell RNA paired with T cell receptor (TCR) sequencing. T cells were found as the dominant immune cell infiltration. Intriguingly, a majority of CD4^+^ in T cell infiltrates was only observed in VKHD but not in BD. In agreement with this discrepancy between VKHD and BD, T cell clonality analysis and Clonotype Neighbour Graph Analysis (CoNGA) also revealed a selective expansion of pro-inflammatory CD4^+^ T cell clones in VKHD. In contrast, BD showed a selective expansion of pro-inflammatory CD8^+^ T cell clones. Our study reveals distinct immunopathogenesis of uveitis driven by CD4^+^ T cells in VKHD and by CD8^+^ T cells in BD, and therefore suggests differential approaches for their diagnosis and therapy.

## 1. Introduction

Uveitis is an inflammatory disorder of the uveal tract of the eye that can be caused by multiple reasons. This vision-threatening disease can cause serious complications such as visual impairment and blindness in 35% of patients ^[1]^. Uveitis can be divided into infectious and non-infectious types according to its etiology. As a more common type, non-infectious uveitis is often related to autoimmune diseases, such as Behçet’s disease (BD) and Vogt-Koyanagi-Harada disease (VKHD). Although BD and VKHD show the characteristics of familial aggregation, they differ in geographical distribution. BD is more prevalent in Asia, the Middle East and the Mediterranean than other regions and is characterised by three main clinical features of nongranulomatous uveitis, oral ulcers and genital ulcers ^[2]^. VKHD is considered to distribute worldwide, with a higher occurrence in people with darker skin ^[3]^. The clinical features of VKHD are bilateral granulomatous pancreatitis, polio, vitiligo, and central nervous system abnormalities ^[4]^.

The immunological pathogenesis of BD and VKHD remains elusive although the dysregulated function of T cells, including cytotoxic CD8^+^ T cells or CD4^+^ T cell subset Th1 and Th17 cells, have been implicated in the development of both BD and VKHD ^[5, 6]^. To gain insight into the immunological pathogenesis of human uveitis, we decided to perform single-cell RNA sequencing (scRNA-seq) of cellular infiltrates in aqueous humour from uveitis patients. Due to the significant involvement of T cells in immunopathogenesis, we included T cell receptor (TCR) sequencing. We discovered that uveitis in VKHD was associated with a strong clonal expansion of effector CD4^+^ T cells expressing pro-inflammatory cytokines *IFNG* and *TNF*. In contrast, uveitis in BD was associated with a selective clonal expansion of effector CD8+ T cells expressing cytotoxic molecules *GZMB* and *PRF1*. The different immunopathogenesis of non-infectious uveitis distinctively driven by CD4^+^ T cells in VKHD and CD8^+^ T cells in BD strongly suggests differential approaches for the diagnosis and therapy of uveitis in VKHD and BD.

## 2. Material and methods

### 2.1 Resource availability

#### 2.1.1 Lead contact

Further information and requests for resources and reagents should be directed to and will be fulfilled by the corresponding authors Di Yu and Yong Tao.

#### 2.1.2 Data and code availability

The datasets during this study are available at the *Science DataBase*: https://www.scidb.cn/s/bAF7fe. Codes are available at https://github.com/HongjianSun/Uveitis-project.

### 2.2 Experiment model and subject details

This study was performed at Beijing Chaoyang Hospital, Capital Medical University, Beijing, China. The ethics approval was obtained from the Ethics Committee of Beijing Chaoyang Hospital. Written informed consent was obtained from all patients. Two VKHD patients and one BD patient (30-40 years old, > 3 years of disease duration) were enrolled at the Ophthalmology Department of Beijing Chaoyang Hospital affiliated with Capital Medical University. All patients had bilateral or unilateral vision loss and had cataract surgeries.

### 2.3 Method details

#### 2.3.1 Single-cell collection

Aqueous humour of about 100 μL was taken from individual patients after an anterior chamber paracentesis (ACP). The ACP was carried out by using a 30-gauge needle, on a 1 mL insulin syringe, via the temporal limbal approach and a rolling technique. Aqueous humour was aspirated and immediately aliquoted into a microfuge tube. All the aqueous humour samples were centrifuged at 800 g for 15 minutes. After removing the supernatant, the cells were suspended in red blood cell lysis buffer (Solarbio) and incubated on ice for 2 min to lyse red blood cells. The pellets were washed twice with PBS.

#### 2.3.2 Single-cell RNA sequencing

10X Genomics Platform was used for single-cell RNA sequencing. Single-cell suspension with gel beads (containing the pre-made 10X primers) and Master Mix together formed Gel Bead-In-Emulsions (GEMs). Cleavage and reverse transcription were performed in GEMs. After cDNA amplification was completed, we performed a quality inspection and finally constructed the library. The Illumina Novaseq 6000 sequencing platform was used for sequencing, obtaining sequencing data, and performing subsequent data analysis.

Fastq reads were initially filtered using *Trimmomatic*^[7]^. Next, we used Feature Counts software to quantify the expression of each gene, and counts were obtained for each sample. In our analysis, a qualified gene expression profile from a single cell should have one or more counts in a sample. The expression level of each gene was converted to a transcript per million (TPM) value. Then, the log-normalized expression values were transformed.

#### 2.3.3 Data processing

*Seurat*^[8]^ was used for scRNA-seq data processing. *Doubletfinder* R package was used to identify doublets^[9]^ and 7.5% was removed as the recommendation. Subsequently, cells from three patient samples were integrated using the *IntegrateData* package to eliminate batch effects.

#### 2.3.4 Data analyzing

*The data were normalized and scaled by NormalizeData* and *ScaleData* in *Seurat*. Unsupervised clustering was performed using *FindClusters* in *Seurat* by a shared nearest neighbour modularity optimization.

Differential gene expression analysis was performed using *FindMarkers* and *FindAllMarkers* in *Seurat* with the parameter of adjusted P-value threshold<0.05. Trajectory analysis was performed by *learn_graph, order_cells* and *plot_cells* in *Monocle3* R package and based on UMAP reduction method^[10]^.

#### 2.3.5 Visualization

scRNA-seq data were visualized using the dimensionality reduction method of UMAP (Uniform Manifold Approximation and Projection). Stacked violin plots were made by a modified function of *Vlnplot* in *Seurat*. Heat map showing the top 5 highest differential expressed genes in each cluster was made by *DoHeatmap* function.

#### 2.3.6 Enrichment Analysis

Gene set enrichment analysis (*GSEA*) was performed^[11]^ with gene sets from *Molecular Signatures Database* v7.4. The enrichment scores (ES) for a gene set reflect the extent to which a transcriptome is overrepresented at the top or bottom of a sequence list of genes, while Normalized Enrichment Scores (NES) are used for comparing the enrichment results through the gene sets. The P-value and the False Discovery Rate (FDR) are both parameters indicating the confidence level of gene set enrichment.

#### 2.3.7 V(D)J sequencing

V(D)J sequencing were analyzed by *CellRanger*. The *CellRanger* VDJ function was used for Contig assembly, cross-cell Consensus sequence, CDR3 Clonotype typing of Loupe V(D)J Browser analysis results on TCR gene V(D)J region sequences of a single cell. *Seurat* and *scRepertoire*^[12]^ were used to add VDJ sequencing data as additional data to merge with single-cell sequencing data.

#### 2.3.8 Clonotype neighbour graph analysis

Clonotype neighbour graph analysis(*CoNGA*) identifies the correlation between T cell gene expression(GEX) and TCR sequence by constructing a GEX similarity graph based on the gene expression, and a TCR sequence similarity graph based on *TCRdist* for each clonotypes’ CDR3 sequence, and then find a statistically significant overlap between them. Overlap is assessed on a score that reflects the likelihood of seeing equal or greater overlap by chance, named the CoNGA score. The more the overlap between GEX and TCR graph, the smaller the CONGA score will be. Then, Clonotypes with CoNGA scores below a threshold are grouped based on shared GEX and TCR cluster assignments into CoNGA clusters. Clonotypes within each CoNGA cluster carry their initial GEX and TCR cluster identities, which are combined together and used as a group ID for the CoNGA cluster.

#### 2.3.9 Statistical analysis

The *Circos* ^[13]^ was created to illustrate the correlations between two groups. Pie charts and histograms were made by *Prism V9* and *Microsoft Excel*.

## 3. Result

### 3.1 T cells are the dominant immune cell type infiltrating in uveitis lesions in both BD and VKHD

Three uveitis patients at the age of 30-40 years old, one diagnosed with BD and two diagnosed with chronic recurrent VKHD volunteered to participate in this study. They all had decreased vision and had cataract surgery more than two years before the sample collection in 2019 (**Fig. 1a**). We choose 5’ whole transcriptome gene expression (10×Genomics) to gain the information for both transcriptomes and paired full-length TCR for immune repertoire analysis. The library preparation, sequencing, read mapping, gene count normalization and doublet removal were conducted using a standard pipeline (**Fig. 1b**) ^[14]^.

**Figure 1.**
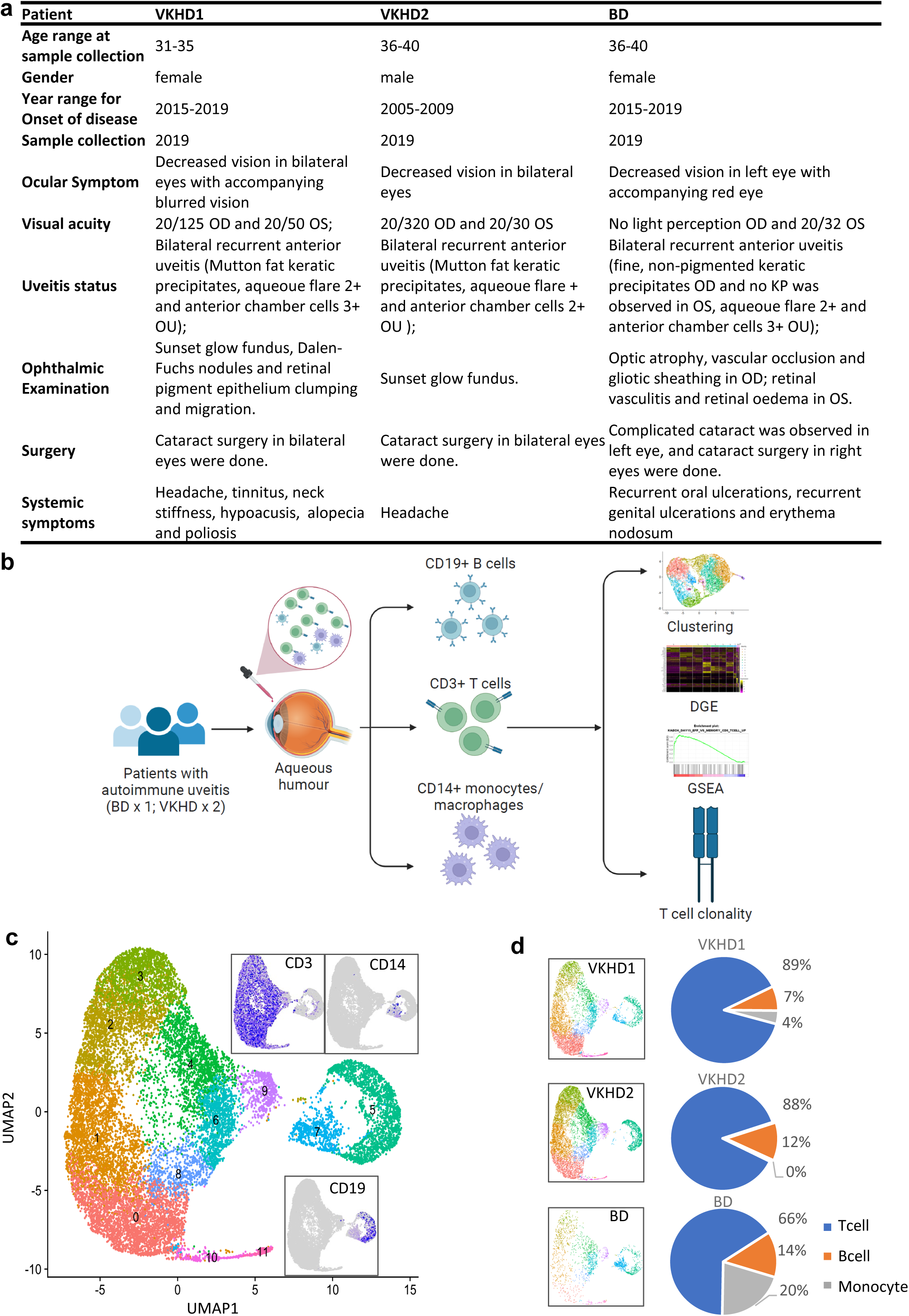
T cells are the dominant immune cell type infiltrating in lesions of non-infectious uveitis. **(a)** Demographic and clinical information of VKHD and BD patients. **(b)** Schematics of the research design. **(c)** UMAP plot showing unsupervised clustering of cells in aqueous humour analysed by scRNA-seq (pooled data from three patients); subgraphs showing the gene expression of *CD3, CD14* and *CD19*. **(d)** UMAP plots showing single cells and clusters in samples from each patient sample (left panels) and frequencies of CD3^+^ T cells, CD19^+^ B cells and CD14^+^ monocytes (right panels).

After the integration of data from three samples, a total of 17,961 cells underwent the downstream analysis. After doublet removing and data integration, the single cells were visualized by UMAP in two-dimensional space. Using KNN method for clustering, the immune cells in uveitis were classified into 12 clusters (**Fig. 1c, S1a**). *CD3*^+^ T cells (cluster 0-4, 6, 8-11) were the dominant immune cell population (11,983 cells) accounting for 44.6% (856/1919), 72.7% (4557/6268) and 67.2% (6570/9774) of total cells from BD, VKHD1 and VKHD2 patients. *CD19*^+^ B cells (cluster 5) were approximately 10% in all samples. Interestingly, *CD14*^+^ monocytes (cluster 7) showed a higher percentage in BD (20.6%) than those in VKHD (VKHD1: 4.0%; VKHD2: 0.4%) (**Fig. 1d, S1b**).

### 3.2 T cells in uveitis lesions are composed of distinct functional subsets

We then focused on T cells as the major immune cell population and identified 11 clusters (**Fig. 2a**), which were predominantly αβ T cells (**Fig. 2b**). We didn’t observe significant enrichment of genes for TCR d and γ, Vα24/Vβ11 associated with natural killer T (NKT) cells, or Vα2/Vα7/Vβ2/13 associated with mucosal-associated invariant T (MAIT) cells in particular subsets ^[15, 16]^ (**Fig.S2a**). Clusters 1, 2, 4, 5, 7, 8, 9 and 10 expressed *CD4* while clusters 3 and 6 expressed *CD8A* and *CD8B*. Cluster 0 are CD4^-^CD8^-^ (double negative, DN) T cells, a population which is rare in a normal condition but expands in inflammation and autoimmune diseases ^[17]^ (**Fig. 2b**). Notably, VKHD samples showed CD4^+^ T cells as the clear majority (VKHD1: 68%; VKHD2: 57%) with CD4/CD8 ratios as 4.3 and 2.6. In contrast, CD4^+^ and CD8^+^ T cell numbers were largely comparable in BD (**Fig. 2c**), with the CD4/CD8 ratio of 1.4. This difference suggests a potential difference in immunopathogenesis of non-infectious uveitis of BD and VKHD.

**Figure 2.**
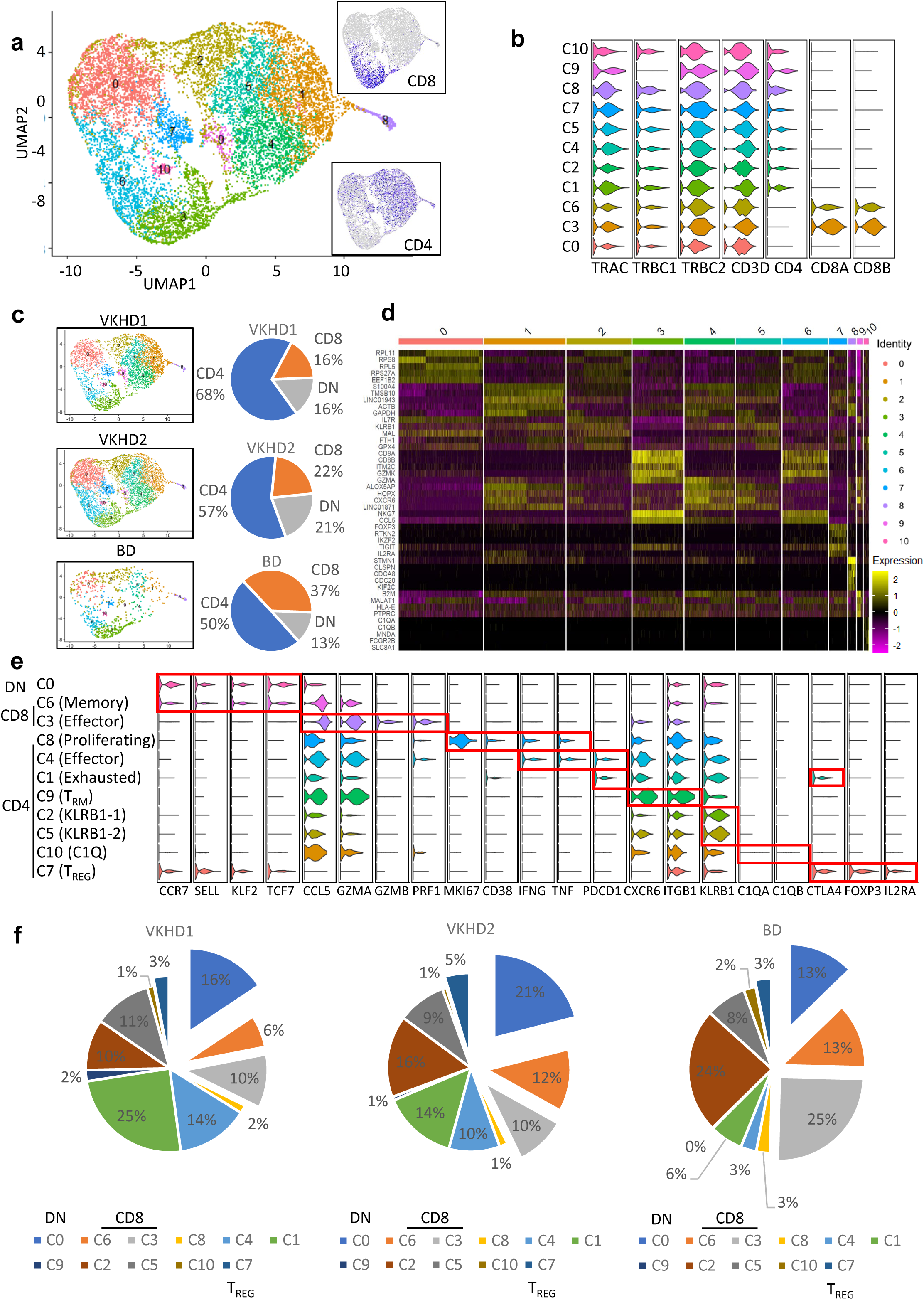
T cell functional subsets in non-infectious uveitis differ between VKHD and BD. **(a)** UMAP plot showing unsupervised clustering of CD3^+^ T cells (pooled from three patients); subgraphs showing the gene expression of *CD4* and *CD8* (both *CD8A* and *CD8B*). **(b)** Violin plots showing the expression of indicated TCR and co-receptor genes in each cluster. **(c)** UMAP plots showing single cells and clusters in samples from each patient (left panels) and frequencies of CD4^+^ T cells, CD8^+^ T cells and CD4^-^CD8^-^ (double negative, DN) T cells (right panels). **(d)** Heatmap showing top 5 differentially expressed genes in each cluster. **(e)** Violin plots of key marker genes for annotating clusters, with the annotation of clusters to the left. **(f)** Pie charts showing the frequencies of each T cell cluster in samples from individual patients.

Using differential gene expressions (DGE) among different clusters (**Fig. 2d**), we annotated their states and functions. CD4^-^CD8^-^ T cells (Cluster 0, C0) were resting T cells with low expression of effector molecules (**Fig. 2e**). Within the CD8^+^ T cells, C6 was a central memory population with the higher expression of the chemotaxis and trafficking receptor genes *CCR7* and *SELL* (encoding CD62L) for recirculation, while C3 was the effector subset with significant expression of effector function genes for C-C chemokine ligands (*CCL5*), granzymes (*GZMA* and *GZMB*) and perforin (*PRF1*) (**Fig. 2e**). Gene Set Enrichment Analysis (GSEA) demonstrated that DGE between C3 and C6 was positively associated with gene expression in effector CD8^+^ T cells as compared to memory or naïve CD8^+^ T cells in the model of acute infection (**Fig. S2b**). The trajectory analysis suggested that CD8^+^ T cells underwent a relatively linear differentiation with a likely progression from memory to effector (**Fig. s2c**).

Within the CD4^+^ T cells, C8 showed a high expression of the proliferation marker *MKI67* (encoding KI-67). Such a highly proliferative population also expressed CD38 and proinflammatory effector genes *IFNG* and *TNF* (**Fig. 2e**), which were recently shown to be correlated with disease severity in COVID-19 ^[18]^. Compared to C8, C4 expresses higher amounts of *IFNG* and *TNF*. They reduced proliferation but underwent terminal differentiation with an upregulation of *PDCD1* (encoding PD-1). Notably, C1 showed the highest levels of *PDCD1* and *CTLA4*, suggesting an exhaustion state with the downregulation of effector molecules (**Fig. 2e**). DGE between C4 and C1 was enriched with the signatures of T cell exhaustion by GSEA (**Fig. S2d**).

C9 is tissue-residential memory T (T_RM_) cells with the expression of chemokine receptor and integrin *CXCR6 and IGTB1*. C2 and C5 express a high level of inhibitory receptor *KLRB1* (encoding CD161). C10 upregulated the initiator of the complement classical pathway *C1QA* and *C1QB*, which were reported to restrain autoimmune response ^[19]^. C7 was regulatory T (T_REG_) cells with the high expression of markers *CTLA4, FOXP3* and *IL2RA* (encoding CD25) (**Fig. 2e**). The trajectory analysis of conventional CD4^+^ T cells (C8, CD4, C1, C9, C2, C5 and C10) suggested that recently activated and proliferating CD4^+^ T cells (C8) may further differentiate and branch into effector (C4), exhausted (C1) and KLRB1^+^ (C2 and C5) clusters (**Fig. S2e**).

With the annotation of T cell functional clusters, we noticed major differences in the frequencies of CD4^+^ and CD8^+^ T cell clusters between VKHD and BD whereas the frequencies of DN T cells were largely comparable among these patients. In agreement with the selectively high CD4/CD8 ratios in VKHD patients, effector (C4) and exhausted (C1) CD4^+^ T cells were in much larger amounts in VKHD but not in BD patients. In contrast, effector (C3) CD8+ T cells were particularly expanded in the BD patient (**Fig. 2f**). These results suggest a notion that uveitis in VKHD and BD is preferentially associated with effector CD4^+^ T cells and effector CD8^+^ T cells, respectively.

### 3.3 Divergent T cell clonal expansion in BD and VKHD uveitis

The antigen is recognised by a specific TCR sequence and its stimulation critically drives T cell proliferation and functional differentiation. We then analysed TCR sequences of T cells in all three patient samples, with an emphasis on clonality by quantifying the frequencies of T cells with a common CDR3 sequence. According to the percentages of the T cell numbers of each T cell clone in the total T cells from a given sample, significantly expanded clones (>0.2% of total T cells) were classified into three groups: Large (> 5% of total T cells), medium (1∼5%), small (0.2-1%) clonotypes. Notably, large clonal types are enriched in effector CD8^+^ T cells (C3) and effector (C4) and exhausted (C1) CD4^+^ T cells (**Fig. 3a**), indicating that effector T cells-mediated inflammation in uveitis is driven by antigen stimulation. Intriguingly, there was no large or medium clonal expansion for Treg cells (C7).

**Figure 3.**
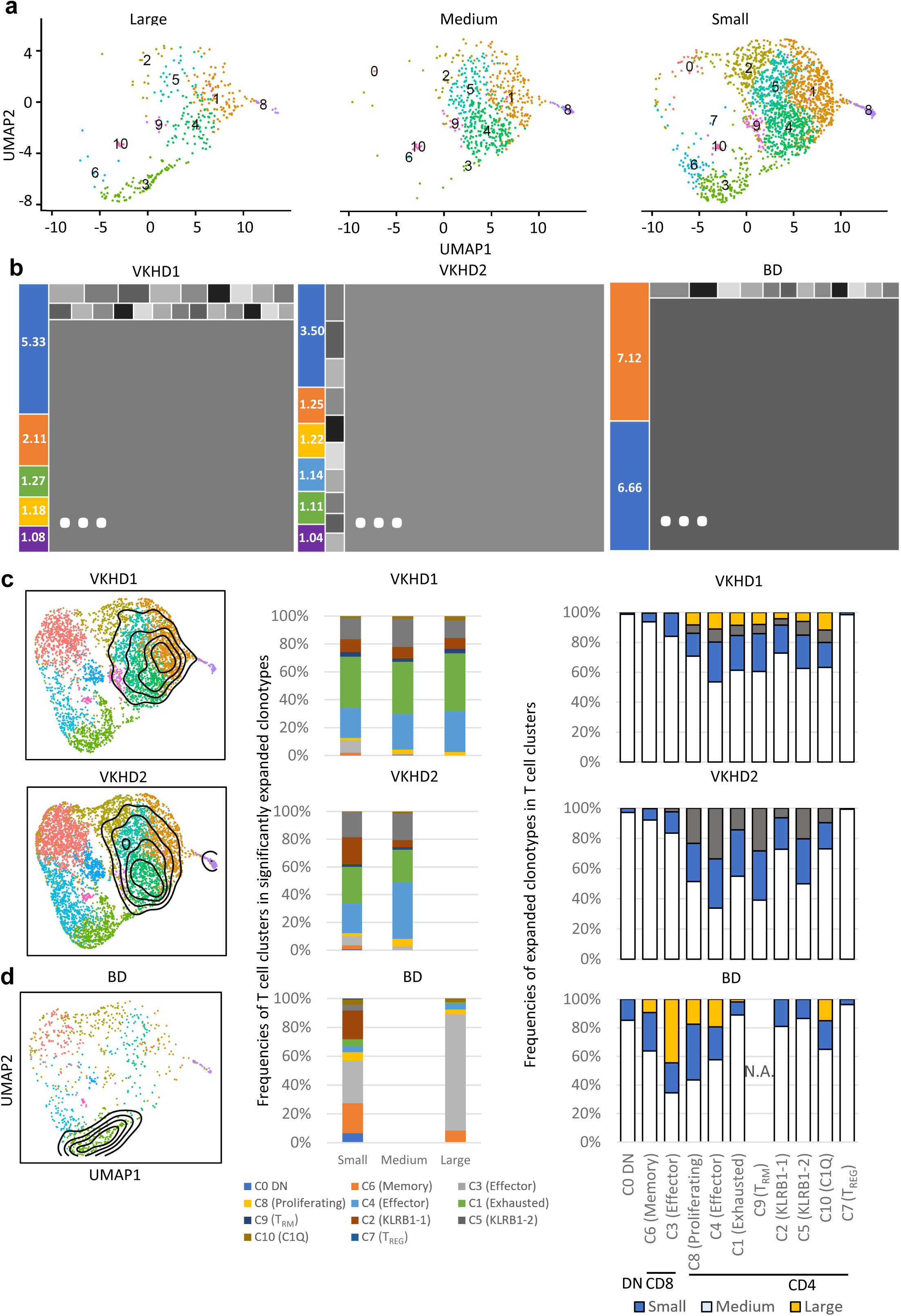
Divergent T cell clonal expansion in BD and VKHD uveitis. **(a)** UMAP plots visualising T cell clonal expansion (pooled data from three patients). *Large*: clonotypes > 5 % of total T cells from the same sample; *Medium*: clonotypes 1-5 % of total T cells from the same sample. *Small*: 0.2-1 % of total T cells from the same sample. **(b)** Treemap plots showing the frequencies of large and medium clonotypes (>1%) in each patient sample. **(c, d)** Contour plots overlayed with UMAP plots showing the distribution of T cells in large and medium clonotypes (left), frequencies of T cell clusters in each category of significantly expanded clonotypes (middle) and frequencies of significantly expanded clonotypes in each T cell cluster (right) for VKHD (c) and BD (d) patients. N.A., not available.

A total of 13 large and medium clonaltypes (>1% of total T cells) were detected in VKHD and BD samples, including 5 in VKHD1, 6 in VKHD2 and 2 in BD (**Fig. 3b, S3**). There were no identical CDR3 sequences among clonotypes from different samples. We then generated topographic maps to visualise the distribution of cells from large and medium clonotypes in each sample. Furthermore, to understand the relationship between clonal expansion and functional T cell clusters, we calculated the frequencies of large, medium and small clonotypes in each functional T cell cluster and also the frequencies of each cluster in large, medium or small clonotypes.

In VKHD, T cell clonal expansions were centred by effector (C4) and exhausted (C1) CD4^+^ T cells (left panels, **Fig. 3c**). Indeed, effector (C4) and exhausted (C1) CD4^+^ T cells accounted for about 70% large and medium clonotypes (middle panels, **Fig. 3c**). Conversely, large and medium clonotypes only accounted for 10-20% of total effector (C4) and exhausted (C1) CD4^+^ T cells (right panels, **Fig. 3c**). The inclusion of small clonotypes could increase the frequencies to about 50%. These results suggest a key role of (auto)antigen-driven CD4^+^ T cell activation in the immunopathogenesis in VKHD.

In BD, T cell clonal expansion was exclusively in effector (C3) CD8^+^ T cells (left panel, **Fig. 3d**). Strikingly, effector (C3) CD8^+^ T cells accounted for about 80% of large clonotypes (middle panel, **Fig. 3d**). Conversely, large clonotypes accounted for >40% of total effector (C3) CD8^+^ T cells (right panel, **Fig. 3d**). These results strongly argue an essential role of (auto)antigen-driven CD8^+^ T cell activation in the immunopathogenesis in BD, which is distinct from VKHD.

In both VKHD and BD, T_REG_ cells (C7) only accounted for a small fraction of T cells (3-5%, **Fig. 2f**). A striking observation was that T_REG_ cells (C7) underwent the least clonal expansion in both VKHD and BD, suggesting that insufficient expansion of T_REG_ cells might posit in the root of losing immune tolerance in non-infectious uveitis. Our previous clinical trials of low-dose IL-2 therapy in systems lupus erythematosus and Sjogren’s syndrome have demonstrated that it could enhance antigen-independent expansion of T_REG_ cells ^[20, 21]^, thus providing a strategy to improve the poor clonal expansion of T_REG_ cells in non-infectious uveitis.

### 3.4 CoNGA identifies similar clonotypes with minor contribution to the inflammation in uveitis

About 50% of effector (C4) and exhausted (C1) CD4^+^ T cells in VKHD (right panels, **Fig. 3c**) and about 40% of effector (C3) CD8^+^ T cells in BD (right panels, **Fig. 3d**) didn’t carry significantly expanded TCR sequences, e.g. not in large, medium or small clonaltypes. This may be explained by TCR-independent activation of T cells, a phenomenon termed ‘bystander activation’ that has been reported in infection and inflammation ^[22]^. An alternative explanation could be that (auto)antigen can activate T cells with similar TCR but such related clonotypes were not identified by identical CDR3 sequences. To test the latter possibility, we applied Clonotype neighbour graph analysis (*CoNGA*), which is a latest method for identifying related clonotypes with similarities in both gene expression and TCR sequences ^[23]^ (**Fig. 4a**).

**Figure 4.**
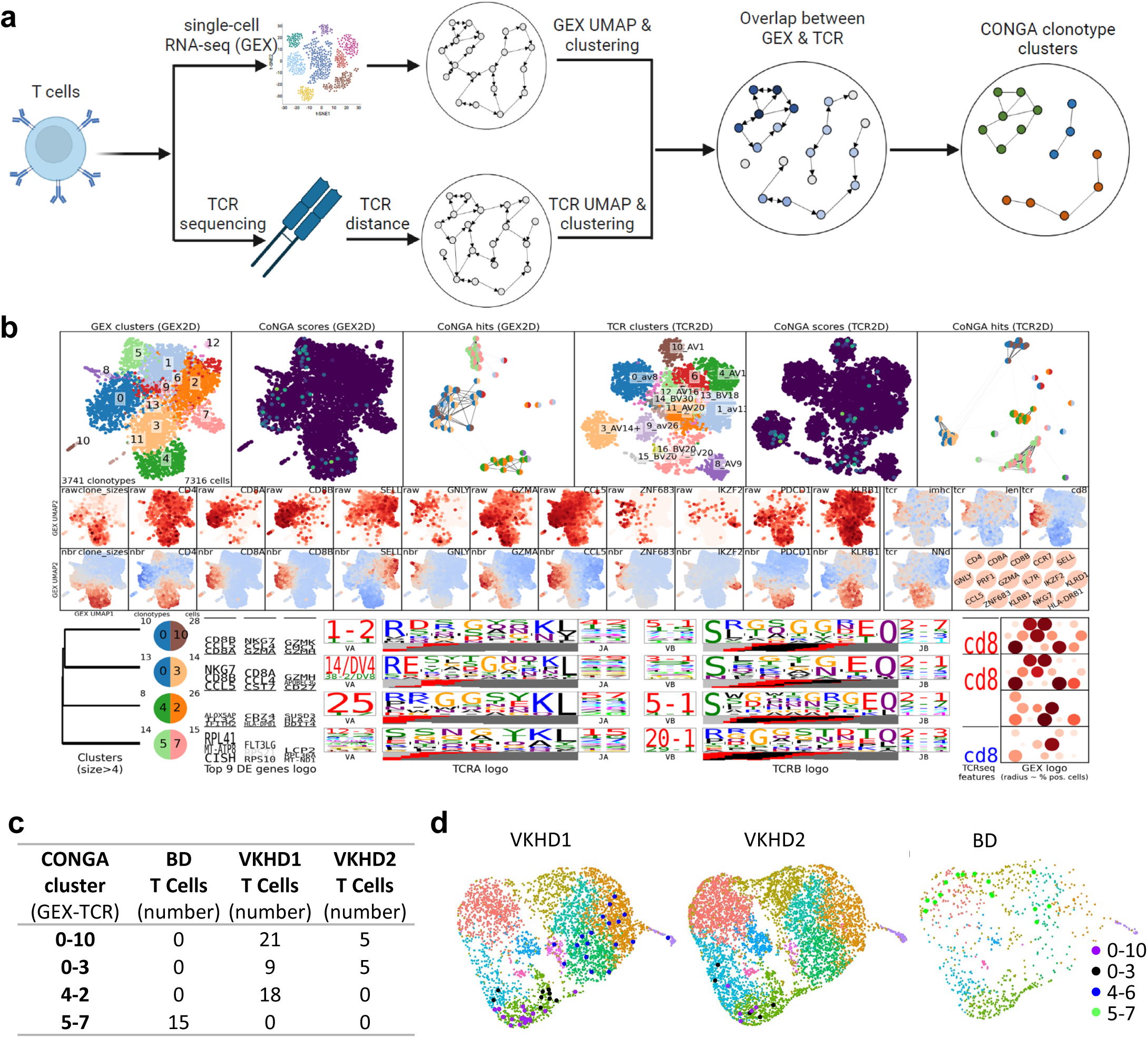
Similar T cell clonotypes in non-infectious uveitis. **(a)** Schematics of clonotype neighbour graph analysis (CoNGA). **(b)** (Top) CoNGA graph-graph relationship plots showing GEX (gene expression) & TCR in pooled data from three patients and unveiling clonotype clusters with similar GEX and TCR. Each dot represents one clonotype. The GEX plot was generated from the gene expression of representative cells from each clonotype. The TCR plot was generated from the TCR distance for each clonotype. (Middle), the expression of feature genes. (Bottom), DEG and TCR sequence logos showing V and J gene usage and CDR3 sequences. DEG and TCR sequence logos are scaled by the adjusted P value of the associations. DEGs with fold-changes less than 2 are shown in grey. **(c)** Table showing the numbers of T cells belonging to the four biggest CONGA clusters. The CoNGA cluster name was based on its GEX (left) and TCR (right) clusters with a dash in the middle. **(d)** UMAP plots showing T cells identified in the top 4 biggest CoNGA clusters in each patient sample. T cells belonging to individual CoNGA clusters were highlighted in different colours.

We performed *CoNGA* on merged single-cell and TCR sequence data. Noticeably, *CoNGA* transformed the scRNA-seq and TCR sequence data and reduced multiple T cells with identical TCR to one cell for *CoNGA* plots ^[23]^. *CoNGA* identified four *CoNGA* clonotypes/clusters (similar gene expression and similar TCR sequences) (**Fig. 4b**). Intriguingly, *CoNGA* clonotypes/clusters 0-10 (GEX Cluster0-TCR Cluster10, **Fig. 4b**) and 0-3 were composed of cells from both VKHD1 and VKHD2. *CoNGA* clonotypes/clusters 4-2 and 5-7 were composed of cells solely from VKHD1 and BD respectively (**Fig. 4c**). We labelled cells identified by *CoNGA* clonotypes/clusters in the UMAP plot and found the majority of such cells were effector (C4) and exhausted (C1) CD4^+^ T cells, or effector (C3) CD8^+^ T cells, except for some cells in the *CoNGA* clonotype/cluster 5-7 belowed to DN (C0) T cells (**Fig. 4d**).

These results suggest that (auto)antigen can activate CD4^+^ or CD8^+^ T cells with similar rather than identical TCR to generate effector cells. We next calculated the contribution of ‘similar’ clonotypes and they only very modestly increased the clonotype frequencies in clusters **(Fig. S4)**. Taken together, ‘similar’ clonotypes can contribute to the immunopathogenesis of uveitis, but ‘identical’ clonotypes appeared to be the major drivers.

## 4. Discussion

Ocular immune privilege has been supported by multiple facets of evidence ^[24]^. The prominent presence of T cell infiltration in non-infectious uveitis suggests they may play a significant role in the immunopathogenesis, which is also supported by results from mouse models of experimental autoimmune uveitis and clinical benefits observed from T cell-targeting therapies ^[25]^. However, the results from human studies were not always consistent, at least partially due to the heterogeneity nature of non-infectious uveitis. Systems immunology approaches have been taken to stratify uveitis patients, such as by peripheral blood transcriptomic analyses ^[26]^.

The latest advancement of scRNA-seq technology allows us to directly investigate immune responses in the ocular microenvironment, thus providing unprecedented opportunities to understand both immunopathogenesis and the heterogeneity among patients ^[27, 28]^.

Our scRNA-seq of cells in aqueous humour from non-infectious uveitis patients confirmed a dominant presence and pro-inflammatory phenotypes of T cells. More importantly, the analyses revealed that T cell phenotypes in VKHD-associated uveitis were different from those in BD-associated uveitis. Uveitis in VKHD patients showed the dominant role of clonally expanded effector CD4^+^ T cells expressing *IFNG* and *TNF*. In contrast, uveitis in the BD patient had few of these effector CD4^+^ T cells but was characterised by clonally expanded effector CD8^+^ T cells expressing *CCL5, GZMA, GZMB* and *PRF1*.

The results strongly suggest the distinctive roles of CD4^+^ and CD8^+^ T cells in the pathogenesis of non-infectious uveitis in VKHD and BD. Such a notion is supported by genetic studies including genome-wide association studies (GWAS). In multiple populations, BD was found associated with MHC class I-specific allele HLA-B*51, while VKHD was associated with MHC class II-specific alleles HLA-DR4/HLA-DRB1 ^[29, 30]^. The important question following our study is what (auto)antigens drive the clonal expansion of effector CD4^+^ T cells in VKHD and the clonal expansion of effector CD8^+^ T cells in BD, with ocular antigens as candidates. If such antigen can be identified to specifically activate individual patient’s T cell clones, the therapy to specifically induce the tolerance to the antigen will be promising as the precedent clinical trial ^[31]^. However, given that identical clonotypes and similar clonotypes identified by CoNGA accounted for only half of effector CD4^+^ and CD8^+^ T cells, our study suggests TCR-independent ‘bystander’ activation of T cells in both VKHD and BD-associated uveitis. This should be taken into account for designing therapeutic strategies for non-infectious uveitis. Another potential application derived from our study is to combine immunological signatures with genetic signatures to stratify patients for therapies that selectively target CD4^+^ or CD8^+^ T cells.

Similar to two recent scRNA-seq studies of aqueous immune cells from 4-5 samples ^[27, 28]^, a major limitation of our study was the limitation of sample numbers. Our results should be validated using a larger sample number. Another limitation was the lack of paired blood samples. If the signatures of expanded T cell clonotypes could be detected in blood samples, a strategy could be considered to monitor blood samples rather than invasive ocular sampling.

## Data Availability

The datasets during this study are available at the Science DataBase: https://www.scidb.cn/s/bAF7fe. Codes are available at https://github.com/HongjianSun/Uveitis-project

## Acknowledgements

This work was supported by the National Natural Science Foundation of China (Grant No.81900849, 82070948, 82071792), Beijing Talent Project (No.2020027), Shunyi District “Beijing Science and technology achievements transformation coordination and service platform” construction fund (SYGX202010) and Capital Health Development Scientific Research Project Grant (to Y.T.). Natural Science Foundation of Shandong Province (Major Basic Program, ZR2020ZD41 to M.S.). Australian National Health Medical Research Council Investigator Fellowship (GNT2009554) and Bellberry-Viertel Senior Medical Research Fellowship (to D.Y.) and The University of Queensland Postgraduate Scholarship (to H.S.).

## Author Contributions

**Hao Kang:** Methodology, Resources, Writing – Original Draft **Hongjian Sun:** Methodology, Software, Formal Analysis, Writing – Original Draft, Visualization **Yang Yang:** Methodology **Minglei Shu:** Supervision **Yunbo Wei:** Methodology **Yu Zhang:** Supervision **Di Yu:** Conceptualization, Writing – Review & Editing, Supervision, Funding Acquisition **Yong Tao:** Conceptualization, Writing – Review & Editing, Supervision, Funding Acquisition

## Additional Information

Supplementary material can be accessed through the online version of this article.

## References

[1] L. Gamalero, G. Simonini, G. Ferrara, S. Polizzi, T. Giani, R. Cimaz. Evidence-Based Treatment for Uveitis. Isr Med Assoc J, 2019;21:475–9.

[2] H. Yazici, E. Seyahi, G. Hatemi, Y. Yazici. Behcet syndrome: a contemporary view. Nat Rev Rheumatol, 2018;14:107–19.

[3] Y. R. Hsu, J. C. Huang, Y. Tao, T. Kaburaki, C. S. Lee, T. C. Lin et al. Noninfectious uveitis in the Asia-Pacific region. Eye (Lond), 2019;33:66–77.

[4] P. Yang, S. Liu, Z. Zhong, L. Du, Z. Ye, W. Zhou et al. Comparison of Clinical Features and Visual Outcome between Sympathetic Ophthalmia and Vogt-Koyanagi-Harada Disease in Chinese Patients. Ophthalmology, 2019;126:1297–305.

[5] L. Du, A. Kijlstra, P. Yang. Vogt-Koyanagi-Harada disease: Novel insights into pathophysiology, diagnosis and treatment. Progress in Retinal and Eye Research, 2016;52:84–111.

[6] B. Tong, X. Liu, J. Xiao, G. Su. Immunopathogenesis of Behcet’s Disease. Frontiers in Immunology, 2019;10.

[7] A. M. Bolger, M. Lohse, B. Usadel. Trimmomatic: a flexible trimmer for Illumina sequence data. Bioinformatics (Oxford, England), 2014;30:2114–20.

[8] Y. Hao, S. Hao, E. Andersen-Nissen, W. M. Mauck, S. Zheng, A. Butler et al. Integrated analysis of multimodal single-cell data. bioRxiv, 2020:2020.10.12.335331.

[9] C. S. McGinnis, L. M. Murrow, Z. J. Gartner. DoubletFinder: Doublet Detection in Single-Cell RNA Sequencing Data Using Artificial Nearest Neighbors. Cell Syst, 2019;8:329-+.

[10] C. Trapnell, D. Cacchiarelli, J. Grimsby, P. Pokharel, S. Li, M. Morse et al. The dynamics and regulators of cell fate decisions are revealed by pseudotemporal ordering of single cells. Nat Biotechnol, 2014;32:381–6.

[11] A. Subramanian, P. Tamayo, V. K. Mootha, S. Mukherjee, B. L. Ebert, M. A. Gillette et al. Gene set enrichment analysis: A knowledge-based approach for interpreting genome-wide expression profiles. Proc Natl Acad Sci U S A, 2005;102:15545–50.

[12] N. Borcherding, N. L. Bormann, G. Kraus. scRepertoire: An R-based toolkit for single-cell immune receptor analysis. F1000Res, 2020;9:47.

[13] M. Krzywinski, J. Schein, I. Birol, J. Connors, R. Gascoyne, D. Horsman et al. Circos: an information aesthetic for comparative genomics. Genome Res, 2009;19:1639–45.

[14] T. Stuart, A. Butler, P. Hoffman, C. Hafemeister, E. Papalexi, W. M. Mauck et al. Comprehensive Integration of Single-Cell Data. Cell, 2019;177:1888-+.

[15] D. G. Pellicci, H.-F. Koay, S. P. Berzins. Thymic development of unconventional T cells: how NKT cells, MAIT cells and γδ T cells emerge. Nature Reviews Immunology, 2020;20:756–70.

[16] H. Y. Greenaway, B. Ng, D. A. Price, D. C. Douek, M. P. Davenport, V. Venturi. NKT and MAIT invariant TCRa sequences can be produced efficiently by VJ gene recombination. Immunobiology, 2013;218:213–24.

[17] D. Brandt, C. M. Hedrich. TCRaβ(+)CD3(+)CD4(-)CD8(-) (double negative) T cells in autoimmunity. Autoimmun Rev, 2018;17:422–30.

[18] Z. Chen, E. John Wherry. T cell responses in patients with COVID-19. Nature Reviews Immunology, 2020;20:529–36.

[19] G. S. Ling, G. Crawford, N. Buang, I. Bartok, K. Tian, N. M. Thielens et al. C1q restrains autoimmunity and viral infection by regulating CD8(+) T cell metabolism. Science, 2018;360:558–63.

[20] J. He, X. Zhang, Y. Wei, X. Sun, Y. Chen, J. Deng et al. Low-dose interleukin-2 treatment selectively modulates CD4(+) T cell subsets in patients with systemic lupus erythematosus. Nat Med, 2016;22:991–3.

[21] J. He, R. Zhang, M. Shao, X. Zhao, M. Miao, J. Chen et al. Efficacy and safety of low-dose IL-2 in the treatment of systemic lupus erythematosus: a randomised, double-blind, placebo-controlled trial. Ann Rheum Dis, 2020;79:141–9.

[22] C. H. Shim, S. Cho, Y. M. Shin, J. M. Choi. Emerging role of bystander T cell activation in autoimmune diseases. BMB Rep, 2022;55:57–64.

[23] S. A. Schattgen, K. Guion, J. C. Crawford, A. Souquette, A. M. Barrio, M. J. T. Stubbington et al. Integrating T cell receptor sequences and transcriptional profiles by clonotype neighbor graph analysis (CoNGA). Nat Biotechnol, 2022;40:54–63.

[24] A. W. Taylor. Ocular immune privilege. Eye, 2009;23:1885–9.

[25] V. L. Perez, R. R. Caspi. Immune mechanisms in inflammatory and degenerative eye disease. Trends Immunol, 2015;36:354–63.

[26] J. T. Rosenbaum, C. A. Harrington, R. P. Searles, S. S. Fei, A. Zaki, S. Arepalli et al. Identifying RNA Biomarkers and Molecular Pathways Involved in Multiple Subtypes of Uveitis. Am J Ophthalmol, 2021;226:226–34.

[27] M. Paley, L. M. Hassman, E. Esaulova, G. L. Paley, J. Laurent, L. Springer et al. Highly expanded clones representing different lymphocyte lineages are present in individual patients with granulomatous uveitis. Investigative Ophthalmology & Visual Science, 2020;61:3661-.

[28] M. Kasper, M. Heming, D. Schafflick, X. Li, T. Lautwein, M. Meyer Zu Horste et al. Intraocular dendritic cells characterize HLA-B27-associated acute anterior uveitis. Elife, 2021;10.

[29] M. Takeuchi, N. Mizuki, S. Ohno. Pathogenesis of Non-Infectious Uveitis Elucidated by Recent Genetic Findings. Frontiers in Immunology, 2021;12.

[30] X. F. Huang, M. A. Brown. Progress in the genetics of uveitis. Genes Immun, 2022;23:57–65.

[31] R. B. Nussenblatt, I. Gery, H. L. Weiner, F. L. Ferris, J. Shiloach, N. Remaley et al. Treatment of uveitis by oral administration of retinal antigens: results of a phase I/II randomized masked trial. Am J Ophthalmol, 1997;123:583–92.

