## Supplementary figures 1-4 for "Single-cell RNA sequencing reveals the distinctive roles of CD4^+^ and CD8^+^ T cells in autoimmune uveitis"

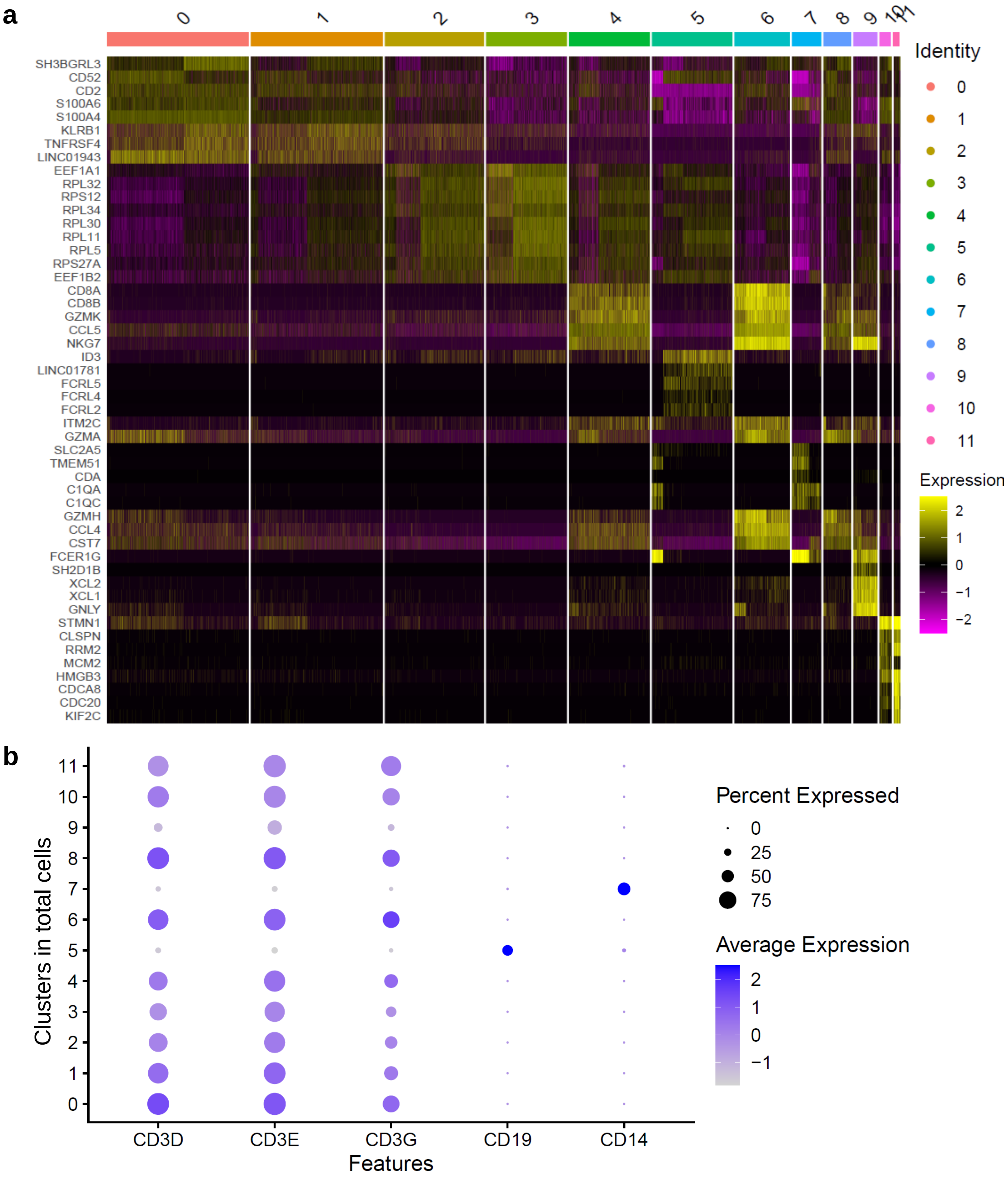


**Supplementary Figure 1**

**(a)** Heatmap for top 5 differentially expressed genes for each clusters of total cells in aqueous humour from non-infectious uveitis patients.

**(b)** The dot plots showing the expression of indicated markers for immune cell types (CD3, CD19, CD14) in each cluster.

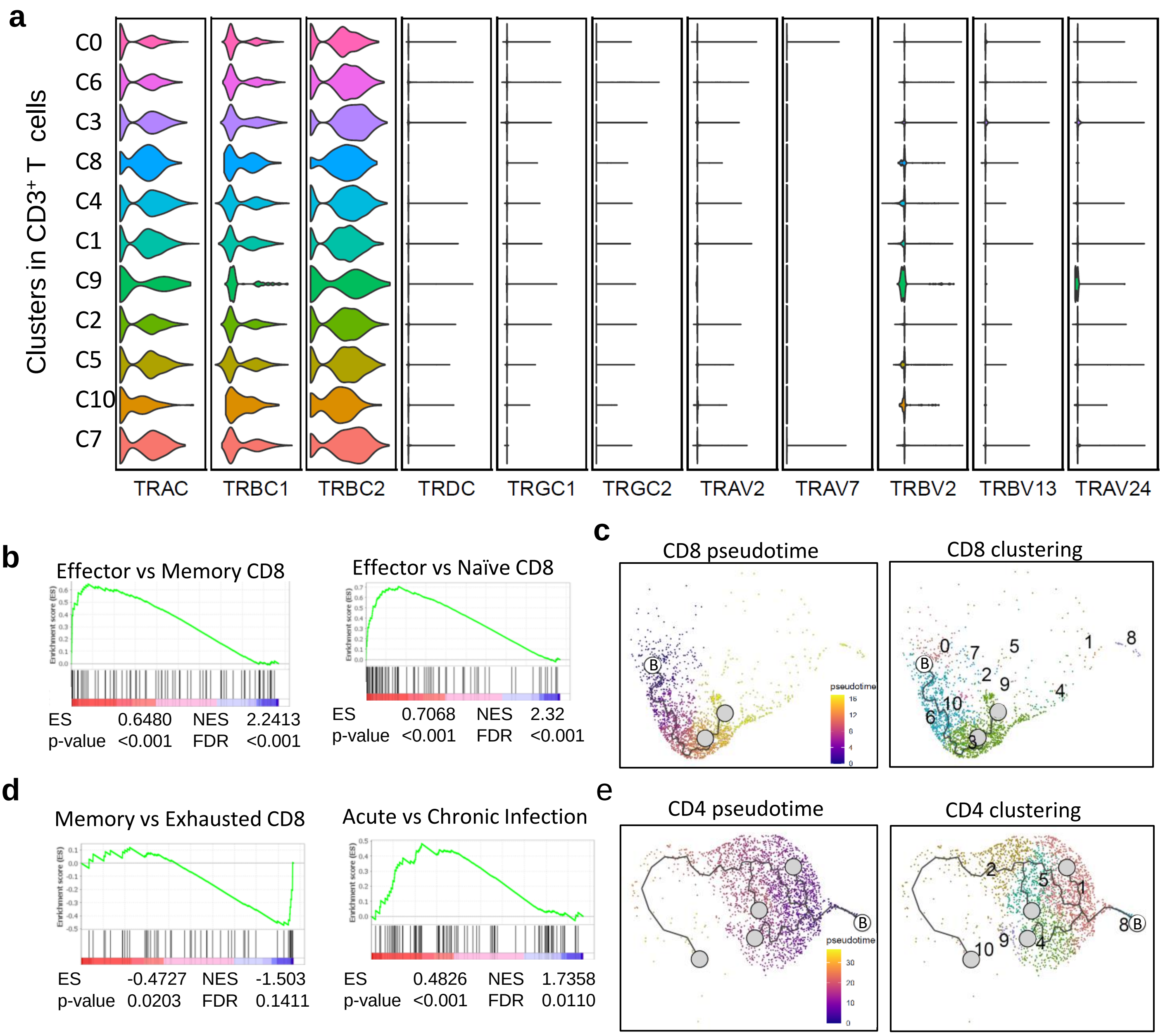


**Supplementary Figure 2**

**(a)** Violin plots showing the expression of TCRs of conventional αβ T cells rather than unconventional γδ T, NKT or MAIT cells.

**(b,d)** GSEA for the DGE for C3 vs C6 (b) and the DGE for C1 vs C4 (d).

**(c,e)** Trajectory analyses of CD8$^{+}$ (c) and conventional CD4$^{+}$ (e) T cells, shown by pseudotime plots (left) and clustering plots (right). Letter B indicates the beginning points of pseudotime analyses whereas grey points indicate the terminal points of pseudotime analyses.

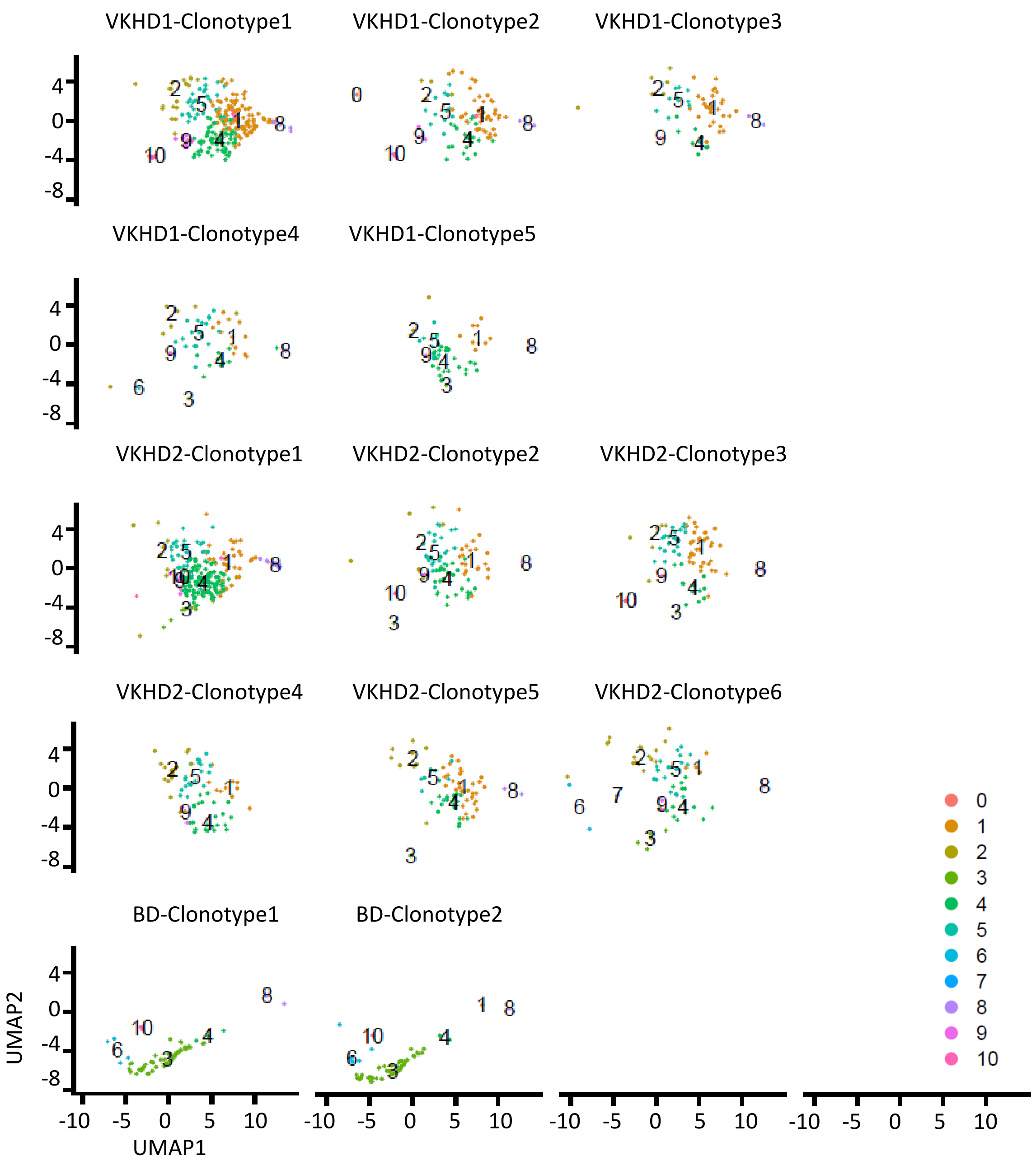


**Supplementary Figure 3**
UMAPs showing expanded clones (>1%) and their distribution among T cell clusters in individual patients (5 clonotypes in VKHD1, 6 clonotypes in VKHD2 and 2 clonotypes in BD).

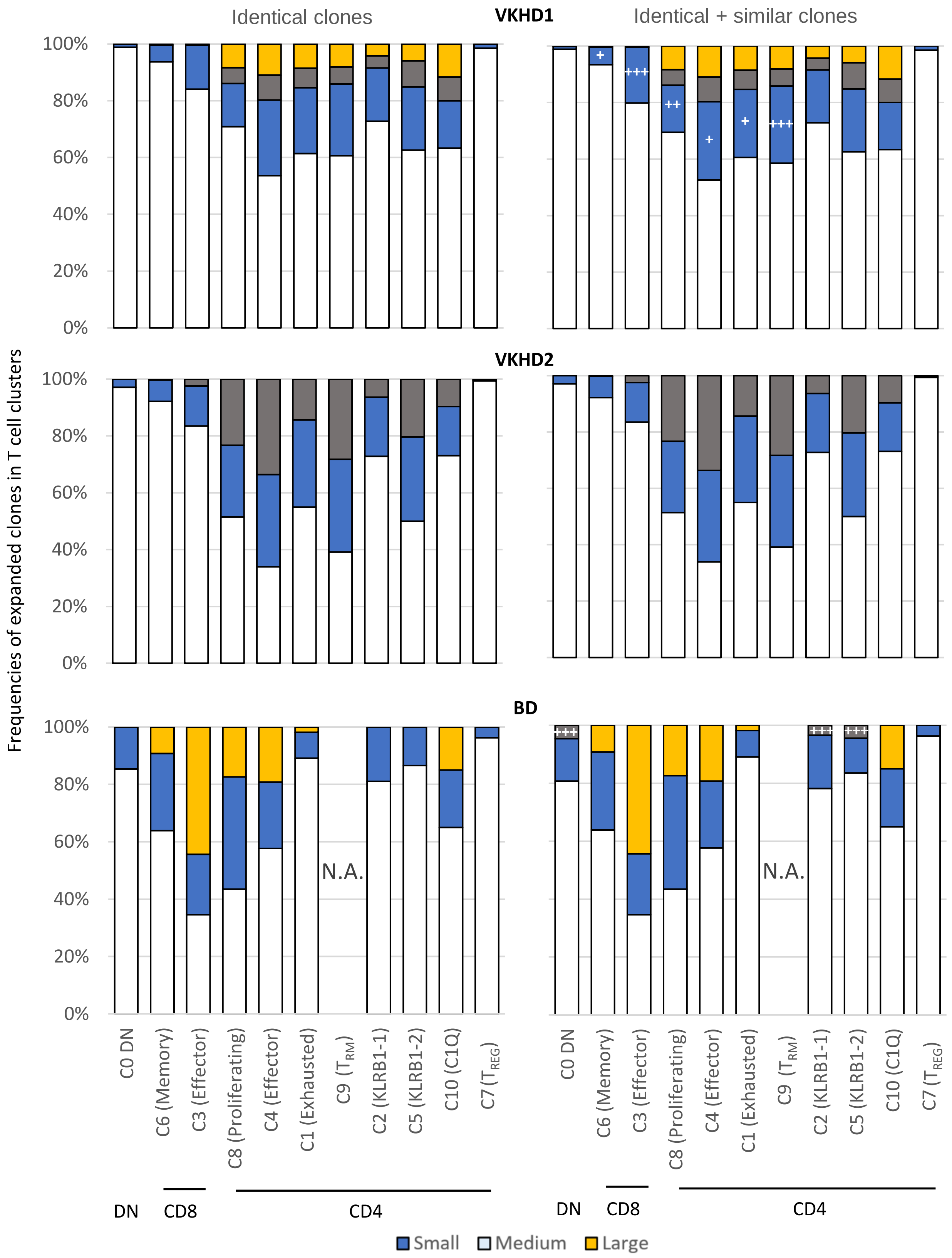


**Supplementary Figure 4**

Statistics of the frequencies of expanded clones in T cell clusters without (left panels) or with (right panels) similar clonotypes identified by CoNGA. The contribution of similar clones were indicated by '+' (<1%), '++' (1-2%) and '+++' (>2%). N.A. means that there are not enough expanded clonal cells available for display.
